# Hypercoagulation detected by Rotational Thromboelastometry predicts mortality in COVID-19: A risk model based on a prospective observational study

**DOI:** 10.1101/2021.04.29.21256241

**Authors:** Lou M. Almskog, Agneta Wikman, Jonas Svensson, Matteo Bottai, Mariann Kotormán, Carl-Magnus Wahlgren, Michael Wanecek, Jan van der Linden, Anna Ågren

**Affiliations:** Department of Anaesthesiology and Intensive Care, Capio St Göran’s Hospital, Stockholm, Sweden; Department of Molecular Medicine and Surgery, Karolinska Institutet, Stockholm, Sweden; Department of Clinical Immunology and Transfusion Medicine, Karolinska University Hospital and Department of CLINTEC, Karolinska Institutet, Stockholm, Sweden; Centre for Psychiatry Research, Department of Clinical Neuroscience, Karolinska Institutet & Stockholm Health Care Services, Region Stockholm, Karolinska University Hospital, Stockholm, Sweden; Division of Biostatistics, Institute of Environmental Medicine, Karolinska Institutet, Stockholm, Sweden; Department of Vascular Surgery, Karolinska University Hospital, Stockholm, Sweden; Department of Physiology and Pharmacology, Karolinska Institutet, Stockholm, Sweden; Perioperative Medicine and Intensive Care, Karolinska University Hospital, Stockholm, Sweden; Coagulation Unit, Hematology Centre, Karolinska University Hospital, Stockholm, Sweden; Department of Clinical Sciences, Danderyd Hospital and Karolinska Institutet, Stockholm, Sweden

**Keywords:** COVID-19, Thromboelastometry, Coagulopathy, Thrombosis

## Abstract

**Background:** Severe disease due to COVID-19 has been shown to be associated with hypercoagulation. Early identification of prothrombotic patients may help guiding anticoagulant treatment and improve survival. The aim of this study was to assess Rotational Thromboelastmetry (ROTEM^®^) as a marker of coagulopathy in hospitalized COVID-19 patients.

**Methods:** This was a prospective, observational study. Patients hospitalized due to a COVID-19 infection were eligible for inclusion. Conventional coagulation tests and ROTEM were taken after hospital admission, and patients were followed for 30 days. Patient characteristics and outcome variables were collected, and a prediction model including variables age, respiratory frequency and ROTEM EXTEM-MCF, was developed using logistic regression to evaluate the probability of death.

**Results:** Out of the 141 patients included, 18 (13%) died within 30 days. D-dimer (p=0.01) and Activated Partial Thromboplastin Time (APTT) (p=0.002) were increased, and ROTEM EXTEM-/INTEM-CT (p<0.001) were prolonged in non-survivors. In the final prediction model, the risk of death within 30 days for a patient hospitalized due to COVID-19 was increased with increased age, respiratory frequency and EXTEM-MCF. Longitudinal ROTEM data in the severely ill subpopulation showed enhanced hypercoagulation. In our in vitro analysis, no heparin effect on EXTEM-CT was observed, supporting a SARS-CoV-2 effect on initiation of coagulation.

**Conclusions:** Here we show that hypercoagulation measured with ROTEM predicts 30-day mortality in COVID-19. Longitudinal ROTEM data strengthen the hypothesis of hypercoagulation as a driver of severe disease in COVID-19. Thus, ROTEM may be a useful tool to assess disease severity in COVID-19, and could potentially guide anticoagulation therapy.

## Background

The global emergence of the corona virus disease 2019 (COVID-19), caused by the severe acute respiratory syndrome coronavirus 2 (SARS-CoV-2), has evolved rapidly achieving pandemic proportions with dire consequences for human health and welfare (1). Though several risk factors for severe disease are known (e.g. high age, obesity, diabetes, chronic pulmonary disease) (2) there is still a need of prognostic models focusing on identifying patients at high risk of death.

Resent reports indicate a high incidence of thrombotic events in COVID-19 patients treated in Intensive Care Units (ICUs) (3), even in patients receiving therapeutic anticoagulation (4). This suggests that an essential pathophysiological component of COVID-19 is related to a widespread and persistent hypercoagulation (5). The underlying mechanisms of the prothrombotic state remains to be clarified (6).

Elevated levels of fibrin degradation products (e.g. D-dimer) have consistently been suggested as a strong prognostic factor associated with poor outcome (7), and early identification of patients at risk of developing thromboembolic complications due to COVID-19 infection may contribute to more adequate antithrombotic strategies. Rotational Thromboelastometry (ROTEM^®^) is a clinically well-established blood test, used for monitoring coagulopathy (8)(9), also in cases where conventional coagulation tests (CCTs) may fail (10)(11). ROTEM variables may be affected earlier during the disease course in COVID-19 compared with other markers (e.g. D-dimer) and may therefore be of greater value as a predictor of disease severity.

Previous studies assessing ROTEM in critically ill COVID-19 patients suggest a procoagulant state (12)(13) and this pattern has also been observed in earlier stages of the COVID-19 disease. However, a prolonged EXTEM Coagulation Time (EXTEM-CT), more pronounced in patients at higher care levels, indicates a prolonged initiation of coagulation in COVID-19 (14).

Several risk stratification tools referring to patients with COVID-19 across different settings and populations have been reported (2) and ROTEM in combination with D-dimer has been verified to predict thromboembolic risks in COVID-19 (4). However, no data evaluating ROTEM as a predictor of mortality have, to our knowledge, yet been published.

In this study we aimed to evaluate several markers of coagulopathy in patients hospitalized due to COVID-19. Specifically, we intended to:

1. Develop and test a pragmatic risk stratification score model, using ROTEM data in combination with other known risk factors, to predict 30-day mortality
2. Assess the longitudinal course of ROTEM test results in severe disease
3. Examine the heparin effect detected by ROTEM, analysed in an in vitro experiment
4. Evaluate the D-Dimer/P-fibrinogen ratio as a marker of thrombotic activity.

## Methods

### Study design

The study was a prospective, observational single-center study. Inclusion criteria were hospitalization due to verified COVID-19 infection and age over 18 years. No exclusion criteria were set up. After inclusion a blood sample was taken and ROTEM analyzed, apart from this nothing differed from the standard care. The ROTEM analyses in this study were performed for research purpose only and ROTEM test results, as opposed to other laboratory test results, were not available for the treating physician and did therefore not affect treatment. All patients were followed up after 30 days when outcomes were registered. The study was approved by the Swedish Ethical Review Authority (D-nr 2020-01875). In the ethical approval consent was waivered in severe cases of COVID-19 disease, where patients due to medical conditions were not able to give their consent.

### Study population

All patients enrolled in the study were included at Capio St Göran’s Hospital, Stockholm, Sweden during a four-month recruitment period (May-August 2020). SARS-CoV-2 infection was verified by either standard polymerase chain reaction (PCR) test (n=129, 91.5%) or computed tomography (CT) scan of the lungs presenting typical SARS-CoV-2 findings in addition to typical clinical symptoms (n=12, 8.5%). Patients were admitted either to regular wards with possibility of low-flow oxygen therapy, or to intermediate wards with more advanced ventilation support; non-invasive ventilation (NIV) or nasal high flow oxygen therapy (NHF), or to the Intensive Care Unit (ICU) where, in addition to NIV/NHF, invasive mechanical ventilation support was available.

### Definitions

In the statistical model, the ROTEM variable EXTEM-MCF was deemed the most suitable candidate as a marker of hypercoagulation, based on a previously published analysis of a subset of the data (14).

Comorbidity in the statistical model was defined as prior diagnosis of either hypertension, diabetes, chronic obstructive pulmonary disease (COPD)/asthma or cardiovascular disease.

Previous thromboembolic disease among included patients was defined as diagnosis of either arterial or venous thrombosis registered in the medical journal at any time prior to presentation of COVID-19 symptoms.

### Laboratory testing

ROTEM and CCTs (D-dimer, P-fibrinogen, Activated Partial Thromboplastin Time (APTT), International Normalized Ratio (INR), Antithrombin and Platelet count), were collected after admission (hospital median day 2, [IQR 1-3]. Apart from these blood tests, nothing in the COVID-19 standard care of included patients was changed including ventilation strategies, medications or routine examinations. To examine the longitudinal course of ROTEM in more severely ill patients, we performed repeated testing at day 5 and day 10 after the first blood test. Patients who were discharged from hospital care prior to the second or third test were not tested. This longitudinal sample will therefore represent the development over time in cases with a more severe disease course. Furthermore, the D-dimer/P-fibrinogen ratio was calculated as a marker of thrombotic activity.

### Anticoagulant therapy

During the study period, anticoagulant therapy was standard of care in COVID-19 pneumonia at Capio St Göran’s hospital and was prescribed according to disease severity and thromboembolic risk profile. Routine anticoagulant treatment administrated after admission was with the Low Molecular Weight Heparin (LMWH) Tinzaparin (Innohep^®^, LEO Pharma, Copenhagen, Denmark) classified after dose regime (low prophylaxis dose (75 IE/kg/24h), high prophylaxis dose (150 IE/kg/24h) or treatment dose (≥175 IE/kg/24h)). Patients with severe disease requiring ventilation support received high prophylaxis doses of antithrombotic treatment, and in case of verified or suspected thromboembolic disease, treatment doses of LMWH were used. Patients in regular wards without risk factors for thromboembolic complications received low-dose prophylaxis.

### ROTEM analysis

ROTEM is an established point-of-care device, used for detecting and monitoring coagulopathy, providing rapid assessment of clot formation to lysis. A ROTEM sigma (Tem Innovations GmbH, Germany) was used for thromboelastometric analyses. Here we present four ROTEM-variables: 1) extrinsically activated assays with tissue factor (EXTEM); (2) intrinsically activated assays using phospholipid and ellagic acid (INTEM); (3) fibrin-based extrinsically activated assays with tissue factor and platelet inhibitor cytochalasin D (FIBTEM) and (4) intrinsically activated assays with the addition of heparinase (HEPTEM). Within every ROTEM-variable, six parameters were quantified: Coagulation Time (CT), which is the time (in seconds) from test start until an amplitude of 2 mm is reached, giving information about coagulation activation/initiation. Clot Formation Time (CFT) corresponds to the time (in seconds) between 2 mm amplitude and 20 mm amplitude, giving information about clot propagation. Maximum Clot Firmness (MCF) is the maximum amplitude (in millimeters) reached during the test, giving information about clot stability. Lysis Index (LI) -30 and LI-60 are the reduction in MCF 30 and 60 minutes after CT, respectively (in percent) (15).

A prolonged EXTEM-CT, short EXTEM-CFT and an increased EXTEM-MCF and/or FIBTEM-MCF suggest a hypercoagulable state with a prolonged initiation of coagulation. A prolonged INTEM-CT compared with HEPTEM-CT illustrates a heparin effect. In order to determine a possible heparin effect on EXTEM-CT, an in vitro analysis was performed where 4 different concentrations of Tinzaparin were added to blood from healthy donors (n=3).

### Statistical analysis

A small number of a priori defined clinical variables were assessed as possible covariates. This list of variables were chosen based on previous knowledge (2), and to be readily available in the clinic: age, respiratory frequency, body mass index (BMI) and comorbidity.

Categorical variables were introduced in the regression model by means of dummy variables. Numeric covariates were transformed with the most suitable power transformation. The transformed variables were entered to the predictive models through natural cubic splines, when significant departures from linearity were detected. The choice of number and location of the knots was based on visual assessment and the Akeike’s information criterion, respectively. The predictive properties of the model were evaluated by calculating the area under the curve (AUC) for the receiver operating characteristic (ROC) curve. Sensitivity, specificity, positive and negative predictive values were calculated. All continuous variables were presented as median and interquartile range (IQR). Two-sided Wilcoxon test was used to test for difference between groups for continuous variables and Fisher’s exact test for categorical data. In the longitudinal data analysis, we used two-sided, paired Wilcoxon test for the ROTEM variables EXTEM-CT, -MCF and -CFT, respectively. P-values below 0.05 were considered statistically significant. Stata statistical software, version 15 (StataCorp LLC) and R, version 3.6.1 was used for statistical analysis and visualizations.

This study was conducted and reported applying the STROBE (Strengthening the Reporting of Observational Studies in Epidemiology) guidelines.

## Results

### Demographic and clinical characteristics

141 COVID-positive patients with a median age of 63 years [IQR 51-75] were included in the study, 87 (62%) were male (Table 1). Comorbidities were common; 45% of patients had a prior diagnosis of hypertension, 24% diabetes, 26% other chronic diseases (renal failure, rheumatological or neurological disease), and 28 patients (20%) previous thromboembolic disease.

**TABLE 1.** Baseline characteristics; survivors, non-survivors and all patients.

|  | Total | Survivors | Non-Survivors | p-value |
| --- | --- | --- | --- | --- |
|  | N=141 | N=123 | N=18 |  |
| Gender male, N (%) | 87 (62%) | 74 (60%) | 13 (72%) | 0.44 |
| Age, years, median (IQR) | 63 (51-75) | 61 (49-74) | 74 (66-80) | 0.002 |
| BMI, kg/m<sup>2</sup>, median (IQR) | 27 (24-31) | 26 (24-30) | 29 (26-33) | 0.04 |
| Previous thromboembolic disease, N(%) | 28 (20%) | 19 (15%) | 9 (50%) | 0.002 |
| Antitrombotic treatment at inclusion, N (%) | 25 (18%) | 16 (13%) | 9 (50%) | <0.001 |
| Diabetes, N (%) | 34 (24%) | 26 (21%) | 8 (44%) | 0.04 |
| Smoking, N (%) | 15 (11%) | 13 (11%) | 2 (11%) | 1 |
| COPD/asthma, N (%) | 20 (14%) | 18 (15%) | 2 (11%) | 1 |
| Hypertension, N (%) | 63 (45%) | 50 (41%) | 13 (72%) | 0.02 |
| Cardiovascular disease, N (%) | 29 (21%) | 19 (15%) | 10 (56%) | <0.001 |
| Malignancy, N (%) | 18 (13%) | 15 (12%) | 3 (17%) | 0.7 |
| Other diseases, N (%) | 37 (26%) | 31 (25%) | 6 (33%) | 0.57 |
| Respiratory frequency at inclusion, breaths/min, median (IQR) | 20 (18-24) | 20 (18-24) | 26 (24-30) | <0.001 |
| Saturation at inclusion, %, median (IQR) | 95 (93-98) | 96 (93-98) | 92 (90-94) | 0.002 |
| Days with COVID symptoms at inclusion, median (IQR) | 10 (7-14) | 9 (7-14) | 11 (9-14) | 0.36 |
| Total hospital days at inclusion, median (IQR) | 2 (1-3) | 2 (1-2) | 3 (2-8) | 0.003 |
| Thrombosis at inclusion, N (%) | 7 (5%) | 6 (5%) | 1 (6%) | 1 |
| Thrombosis after inclusion, N (%) | 8 (6%) | 6 (5%) | 2 (11%) | 0.27 |
| Anticoagulant prophylaxis before ROTEM analysis, N (%) | 101 (72%) | 85 (69%) | 16 (89%) | 0.1 |

IQR=Interquartile Range, BMI=Body Mass Index, COPD=Chronic Obstructive Pulmonary Disease.

Demographic and clinical baseline characteristics for all patients were divided in survivors and non-survivors (Table 1). 18 patients (13%) died within 30 days after inclusion (“non-survivors”). Non-survivors were older and had higher BMI than survivors. Further, among non-survivors, a larger proportion of patients had previous thromboembolic disease, diabetes and hypertension compared with survivors. Non-survivors also had higher respiratory frequency and lower blood oxygen saturation at inclusion compared with survivors. Number of days with COVID-symptoms prior to inclusion did not differ significantly between survivors and non-survivors. Eight patients (6%) had thrombosis during hospitalization (2 pulmonary embolism, 3 myocardial infarction, 3 distal venous thrombosis).

Among non-survivors, 9/18 patients (50%) had ongoing anticoagulant or platelet inhibiting treatment before hospital admission, prescribed prior to their SARS-CoV-2 infection (5 patients were treated with direct oral anticoagulants (DOAC), 1 patient with Warfarin, 1 patient with LMWH, 2 patients with platelet inhibitors). 10 (56%) of non-survivors had cardiovascular disease. Among survivors, 16/123 (13%) had anticoagulant treatment prescribed before inclusion, and 19 (15%) had a history of cardiovascular disease.

### Laboratory test results

Laboratory test results are presented in Table 2. D-dimer and Activated Partial Thromboplastin Time (APTT) were significantly increased in non-survivors compared with survivors. Platelet count, INR, P-fibrinogen and antithrombin did not differ significantly among survivors compared with non-survivors. EXTEM-/INTEM-CT were significantly prolonged in non-survivors compared with survivors. EXTEM-/FIBTEM-MCF were in upper reference ranges and EXTEM-/INTEM-LI30 and -LI60 indicated low fibrinolytic activity in survivors as well as non-survivors.

**TABLE 2.** Laboratory test results at inclusion; survivors, non-survivors and all patients.

|  |  |  |  |  |  |
| --- | --- | --- | --- | --- | --- |
|  | Total | Survivors | Non-Survivors | p-value | Reference range |
|  | N=141 | N=123 | N=18 |  |  |
| Covid-positive, verified, N (%) | 141 (100%) | 123 (100%) | 18 (100%) | - |  |
| D-dimer, mg/L, median (IQR) | 0.7 (0.5-1.4) | 0.7 (0.5-1.3) | 1.6 (0.6-3.0) | 0.01 | < 0.5 |
| Platelet count, 10<sup>9</sup>/L, median (IQR) | 225 (169-291) | 222 (170-289) | 243 (163-345) | 0.49 | men 145-348, women 165-387 |
| APTT, sec, median (IQR) | 26 (24-30) | 26 (24-29) | 31 (27-33) | 0.002 | 24-32 |
| INR, median (IQR) | 1.0 (1.0-1.1) | 1.0 (1.0-1.1) | 1.1 (1.0-1.4) | 0.15 | 0.9-1.2 |
| P-fibrinogen, g/L, median (IQR) | 5.5 (4.4-6.9) | 5.4 (4.3-6.9) | 5.9 (5.3-7.1) | 0.14 | 1.8-3.8 |
| Antithrombin, kIE/L, median (IQR) | 1.0 (0.9-1.1) | 1.0 (0.9-1.1) | 0.9 (0.9-1.1) | 0.18 | 0.8-1.2 |
| D-dimer/P-fibrinogen ratio (%), median (IQR) | 0.015 (0-0.030) | 0.014 (0.001-0.028) | 0.021 (0.001-0.042) | 0.16 |  |
| EXTEM-CT, sec, median (IQR) | 71 (61-87) | 70 (61-84) | 94 (78-146) | <0.001 | 38-79 |
| EXTEM-CFT, sec, median (IQR) | 49 (41-61) | 49 (42-60) | 51 (42-69) | 0.44 | 34-159 |
| EXTEM-MCF, mm, median (IQR) | 71 (67-75) | 70 (67-74) | 74 (69-77) | 0.05 | 50-72 |
| EXTEM-Li30, %, median (IQR) | 100 (100-100) | 100 (100-100) | 100 (100-100) | - | 94-100 |
| EXTEM-Li60, %, median (IQR) | 97 (96-98) | 97 (95-99) | 98 (98-99) | 0.05 | 90-92 |
| INTEM-CT, sec, median (IQR) | 185 (170-199) | 183 (170-195) | 200 (192-216) | 0.003 | 100-240 |
| INTEM-CFT, sec, median (IQR) | 55 (44-69) | 55 (44-70) | 54 (40-63) | 0.41 | 30-110 |
| INTEM-MCF, mm, median (IQR) | 67 (63-72) | 67 (63-72) | 73 (67-74) | 0.02 | 50-72 |
| INTEM-Li30, %, median (IQR) | 100 (100-100) | 100 (100-100) | 100 (100-100) | - | 94-100 |
| INTEM-Li60, %, median (IQR) | 98 (96-100) | 97 (95-99) | 99 (99-99) | 0.05 | 90-93 |
| FIBTEM-MCF, mm, median (IQR) | 29 (24-34) | 28 (24-33) | 30 (28-37) | 0.09 | 9-25 |
| HEPTEM-CT, sec, median (IQR) | 185 (174-198) | 182 (174-196) | 204 (183-213) | 0.01 | 100-240 |

IQR=Interquartile Range, APTT=Activated Partial Thromboplastin Time, INR=International Normalized Ratio.

### Prediction model

The final logistic regression prediction model is:

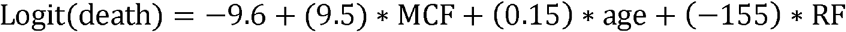

where *Logit(death)* is the predicted log-odds of 30-day mortality (here synonymous with the risk score); MCF is the cubic power of EXTEM-MCF divided by 10^6^; age is in years and not transformed; RF is the reciprocal (1/x) of the respiratory frequency at inclusion.

According to this model, the risk of death within 30 days for a patient hospitalized due to COVID-19 was increased with increased age, respiratory frequency and EXTEM-MCF (all predictors P < 0.05) (*Table 3*).

**TABLE 3.** Logistic regression for mortality at 30 days.

| TABLE 3. Logistic regression for mortality at 30 days. |  |  |  |  |  |  |
| --- | --- | --- | --- | --- | --- | --- |
|  | Coefficient | Standard Error | z | P>\|z\| | [95% Confidence Interval] |  |
| EXTEM-MCF (mm) | 9.49 | 4.65 | 2.04 | 0.041 | 0.37 | 20 |
| Respiratory frequency (breaths/min) | -154,97 | 41.54 | -3,73 | 0.000 | -236,38 | -73,56 |
| Age (years) | 0.15 | 0.047 | 3.27 | 0.001 | 0.061 | 0.25 |
| Constant | -9,59 | 3.89 | -2,47 | 0.014 | -17,22 | -1,97 |

MCF=Maximum Clot Firmness.

The risk score generated by the model may be transformed to the more intuitive variable *probability of death* (Figure 1).

**Figure 1.**
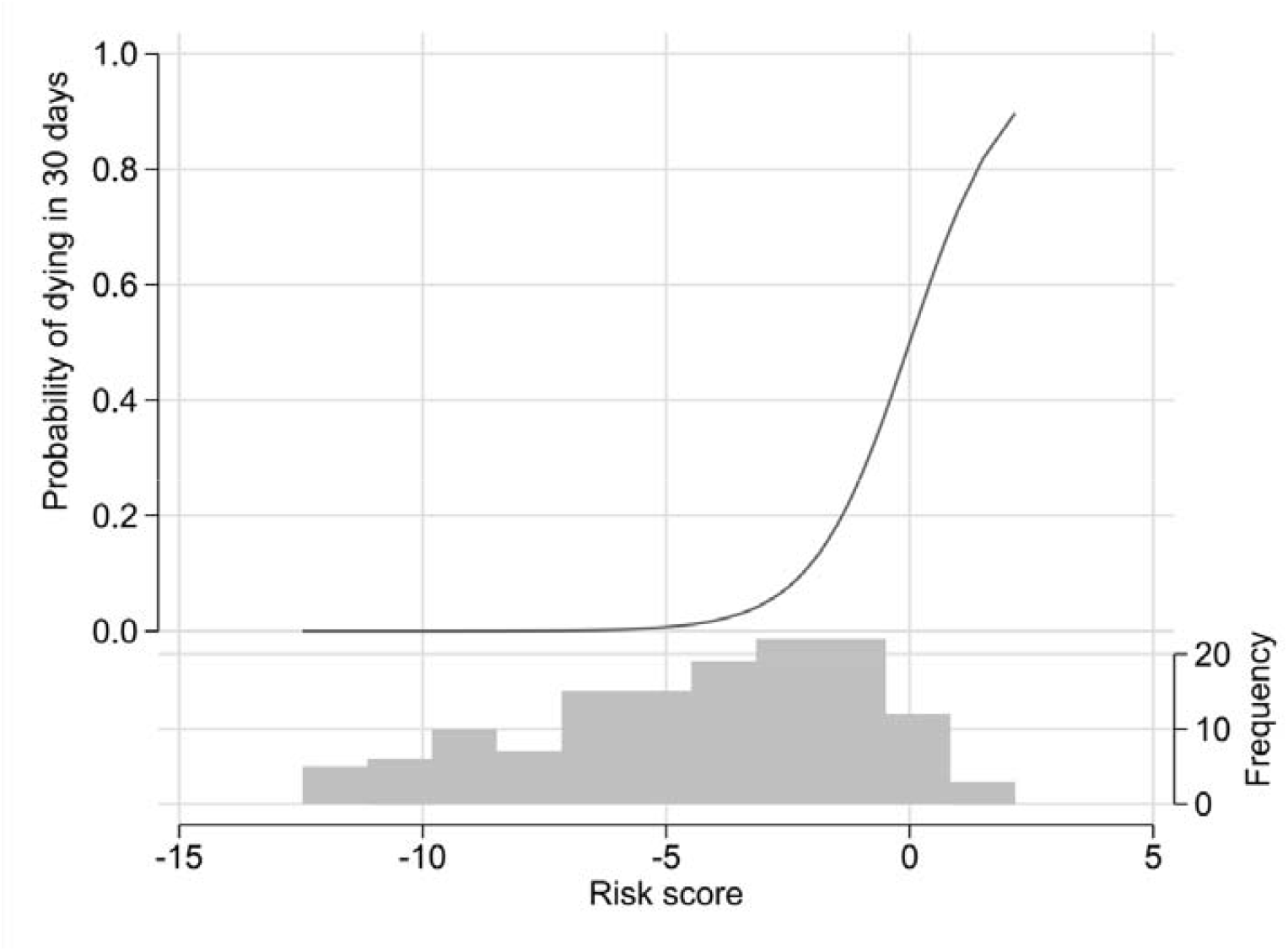
Predicted probability of death vs risk score. Distribution of patients across range of risk scores, corresponding to the logit (logged odds ratio) calculated using logistical regression with the three predictor variables at inclusion (EXTEM-MCF, age and respiratory frequency). The histogram in light gray, presented in the lower part of the figure, shows the distribution of risk scores in the present data (e.g., 12 subjects have a risk score of 0, translating to approximately 50% probability of dying within 30 days). MCF=Maximum Clot Firmness.

Through changing one predictor while keeping other variables constant it is possible to illustrate the effect of the predictors on the probability of death. If, for example, respiratory frequency is kept constant at 20 breaths/min (median value in the full sample), as EXTEM-MCF is increased from 65 to 75, mortality risk increases from 0.1% to 0.4% in a 51-year-old patient [lower age quartile], and from 3.8% to 13.7% in a 75-year-old [higher age quartile] (*Table 4*).

**TABLE 4.** Mortality 30 days in %, related to age (years) and EXTEM-MCF (mm).

| TABLE 4. Mortality 30 days in %, related to age (years) and EXTEM-MCF (mm). | EXTEM-MCF |  |  |
| --- | --- | --- | --- |
|  | 65 | 70 | 75 |
| AGE 51, RF 20 | 0.1% | 0.2% | 0.4% |
| AGE 63, RF 20 | 0.6% | 1.2% | 2.5% |
| AGE 75, RF 20 | 3.8% | 7.0% | 13.7% |

RF=Respiratory Frequency, MCF=Maximum Clot Firmness.

When the model is applied to the data, the ROC curve AUC is 0.91. If the cutoff for probability of death is set to 0.13, this corresponds to a sensitivity of 94%, and specificity 81%, a positive predictive value of 41% and a negative predictive value of 99%.

### Longitudinal data

In the longitudinal analysis, 57 patients were tested a second time, and of these 24 patients were tested a third time. EXTEM-MCF increased from median 73 mm [IQR 65-81] to 76 mm [IQR 68-84] (p<0.001). For the subset of patients tested a third time the value increased from 75.5 mm [IQR 68-83] to 78 mm [IQR 71-85] (p=0.006). EXTEM-CFT decreased from median 48 mm [IQR 31-65] to 44 mm [IQR 32-56] (p<0.001). For the subset of patients tested a third time the second test was 49 mm [IQR 27-71] and the third 46.5 mm [IQR 34.5-58.5] (p=0.13). No significant changes in EXTEM-CT were observed, first test median 82 seconds [IQR 51-113], second test 77 seconds [IQR 52-102] (p=0.19). For the subset tested a third time the second test was 84 seconds [IQR 55-113] and the third 83 seconds [IQR 59-107] (p=0.42), *Figure 2*.

**Figure 2.**
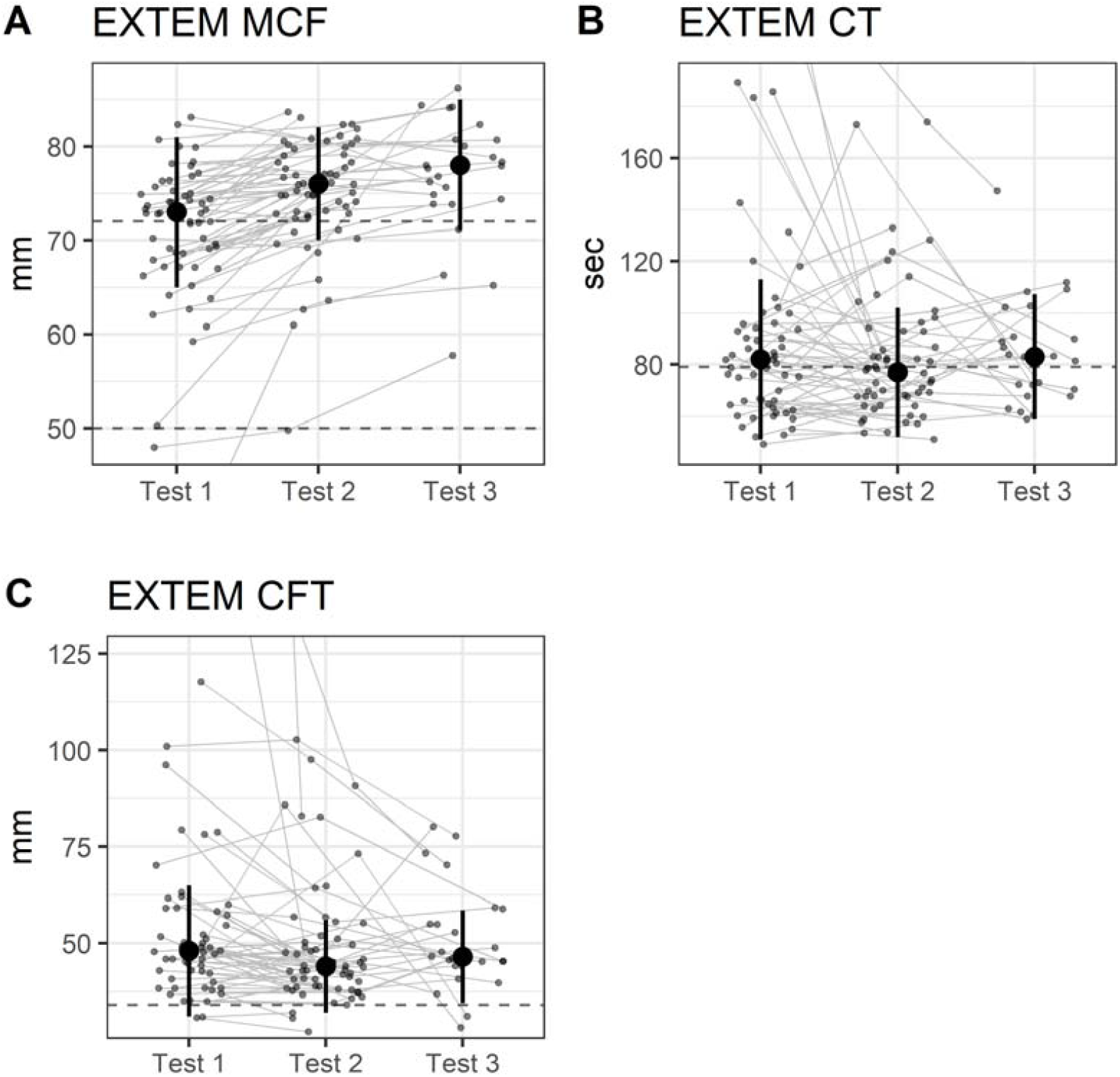
Longitudinal ROTEM data. ROTEM sampled at inclusion, after 5 days and after 10 days. In A) EXTEM-MCF (dashed horizontal lines are upper and lower reference values: 50-72 mm); B) EXTEM-CT (dashed horizontal line is upper reference value: 79 sec); C) EXTEM-CFT (dashed horizontal line is lower reference value: 34 sec). Median values reported with error bars representing IQR. For visualization purposes some outliers are not shown but are represented by lines connecting them to follow up measurements. MCF=Maximum Clot Firmness, CT = Coagulation Time, CFT= Clot Formation Time, IQR= Interquartile Range.

### INTEM-HEPTEM difference

In order to assess the heparin effect on our test results, we evaluated the difference between INTEM-CT and HEPTEM-CT in all patients receiving LMWH before inclusion (N=86). In this subgroup, we did not observe any significant INTEM-HEPTEM CT difference (p = 0.55) indicating no heparin effect on our ROTEM results.

### In vitro analysis

In our dataset, we observed a prolongation of EXTEM-CT, more pronounced in non-survivors compared with survivors. The majority of patients in both groups had received LMWH prior to inclusion (non-survivors 89%, survivors 69%). Previous in-vitro data have shown no effect on EXTEM-CT by LMWH (Dalteparin/Fragmin^®^, Pfizer, New York, USA) in therapeutic doses (16). To determine whether prolonged EXTEM-CT may possibly be associated with a heparin effect at higher doses of LMWH, we performed an experimental study with Tinzaparin in vitro (Table 5) where the results did not indicate any effect on EXTEM-CT with increasing LMWH concentration.

**TABLE 5.** INTEM-/EXTEM-/HEPTEM-CT at different Tinzaparin concentrations.

|  | *Tinzaparin conc* |  |  |  |  |
| --- | --- | --- | --- | --- | --- |
|  | 0 IU/mL | 0.5 IU/mL | 1 IU/mL | 1.5 IU/mL | 2 IU/mL |
| **Donor 1** |  |  |  |  |  |
| INTEM-CT (sec) | 170 | 253 | 368 | 220 | 332 |
| EXTEM-CT (sec) | 43 | 56 | 48 | 49 | 52 |
| HEPTEM-CT (sec) | 172 | 139 | 239 | 148 | 165 |
| **Donor 2** |  |  |  |  |  |
| INTEM-CT (sec) | 231 | 269 | 358 | 447 | 453 |
| EXTEM-CT (sec) | 67 | 60 | 56 | 57 | 60 |
| HEPTEM-CT (sec) | 172 | 180 | 195 | 197 | 218 |
| **Donor 3** |  |  |  |  |  |
| INTEM-CT (sec) | 264 | 353 | 282 | 338 | 348 |
| EXTEM-CT (sec) | 50 | 47 | 49 | 43 | 52 |
| HEPTEM-CT (sec) | 270 | 131 | 249 | 167 | 294 |

### D-dimer/P-fibrinogen ratio

The D-dimer/P-fibrinogen ratio reflects fibrinolysis in relation to fibrin deposition and has previously been defined as a marker of thrombotic activity, where a higher ratio correlates to a more thrombogenic profile (17). In our data, an increased ratio was observed in non-survivors compared with survivors, 0.021 [IQR 0.001-0.042] and 0.014 [IQR 0.001-0.028], respectively (Table 2). The difference was not statistically significant (p=0.16).

## Discussion

This study reports a stratification risk score model where we evaluate the ability of ROTEM to predict mortality in COVID-19 patients. Our results support the concept of an early pronounced hypercoagulability, measured by increased EXTEM-MCF on admission, that is associated with an increased mortality risk. In combination with age and respiratory frequency, which are two easily measured clinical parameters, our model introduces a feasible tool to assess the risk of death in COVID-19 pneumonia.

There is an abundance of findings suggesting that hypercoagulopathy is a crucial component in the pathophysiology of severe COVID-19. Importantly, enhanced anticoagulant treatment has been shown to be associated with reduced mortality (18) as well as a reduction in inflammatory biomarkers (19).

The covariates included in the model (age, respiratory frequency) have in earlier studies been suggested as important predictors of clinical outcome. High age is one of the most frequently reported predictors of poor prognosis in COVID-19 and respiratory frequency has in patients with COVID-19 been described as a predictor of mechanical ventilation and in-hospital mortality (2). When these three predictors were modeled, we observed a high sensitivity of 94% and a high specificity of 81% in our data.

In our patients we typically found prolonged initiation of coagulation (prolonged EXTEM-CT), shortened clot propagation (shortened EXTEM-CFT) with a subsequent forming of a very stable clot with a pronounced clot firmness (increased EXTEM-/FIBTEM-MCF) and coexisting hypofibrinolysis (increased EXTEM-/INTEM-LI60). Our longitudinal analysis supports these findings, where increasing EXTEM-MCF and shortening of EXTEM-CFT were observed confirming enhanced hypercoagulation.

A prolonged EXTEM-CT is, in contrast to other ROTEM variables, not an indication of hypercoagulation. Increased EXTEM-CT is, however, in line with previous studies, where prolongation of prothrombin time with similar activation proteins as in EXTEM, was identified in COVID-19 patients (20)(3)(21). In our in vitro analysis results, no impact on EXTEM-CT was observed with increasing LMWH-doses, supporting the hypothesis that prolonged initiation of coagulation in COVID-19 may be due to viral effects.

Low fibrinolytic activity was observed in survivors as well as non-survivors in our data, indicating hypofibrinolysis/fibrinolysis shutdown in both EXTEM and INTEM (22). Decreased fibrinolysis may partially explain the hypercoagulability observed in COVID-19. Indeed, earlier reports suggest that elevated levels of plasminogen activator inhibitor type 1 (PAI-1), which is one of the most important inhibitors of the fibrinolytic system, may result in lower plasmin activity and hence decreased fibrinolysis. This in turn, may contribute to an imbalance between coagulation and fibrinolysis in COVID-19 (23).

Elevated P-fibrinogen and D-dimer levels have frequently been reported in COVID-19 infected patients and were so also in our data. However, these variables are considered acute inflammatory plasma markers expected to rise during inflammation, and neither parameter has been shown to reliably identify patients with increased thromboembolic risk in COVID-19 (24). Furthermore, earlier data suggest that elevated D-dimer levels in COVID-19 are rather reflecting increased fibrin deposition than increased fibrin breakdown (25).

The D-dimer/P-fibrinogen ratio is an indicator of prothrombotic activity, where a higher ratio suggests a more pronounced thrombotic state. An increased ratio in COVID-19 may reflect the presence of activated coagulation leading to fibrinogen consumption in the pulmonary vasculature, with simultaneous activation of fibrinolysis resulting in elevated d-dimer levels (26). In our data, we observed a higher D-dimer/P-fibrinogen ratio in non-survivors compared with survivors, but the difference was not significant.

Some limitations of this study should be recognized: 1) The sample size was relatively small. 2) Some patients were included somewhat later than the day of admission, which may have reflected test results of different disease stages. 3) Most patients had received antithrombotic treatment prior to inclusion, which may have influenced our lab results. However, given that the ROTEM variable we chose to include as a predictor indicated hypercoagulopathy, we do not presume this created any false positive associations.

The recruitment of patients was performed at a single site with no validation set. The applicability of the model therefore needs to be validated in larger independent cohorts, in order to confirm its generalizability in other settings and populations.

These limitations notwithstanding, we consider our cohort as a representative sample from the first wave of COVID-19 in Stockholm, in which a state of hypercoagulability has been shown to be associated with an increased risk of death. Together, these results indicate that ROTEM is a useful analysing method of coagulopathy in COVID-19 and may be a promising tool to guide anticoagulant treatment.

## Conclusions

In this study, we evaluated ROTEM as a marker of coagulopathy in COVID-19. First, we presented a risk stratification score model where increased EXTEM-MCF, in combination with age and respiratory frequency, were predictive of increased mortality within 30 days. We then assessed the longitudinal disease course where ROTEM supported the hypothesis of enhanced hypercoagulation in severe disease. Our data did not indicate any effect on EXTEM-CT with increasing LMWH concentrations. In conclusion, ROTEM is a potentially useful guide of anticoagulant therapy and suggested as a feasible tool for monitoring disease, which may improve survival in patients with a poor prognosis in COVID-19.

## Data Availability

Shared upon request.

## List of abbreviations

(ROTEM): Rotational Thromboelastmetry
(EXTEM): Extrinsically activated assays with tissue factor
(MCF): Maximum Clot Firmness
(APTT): Activated Partial Thromboplastin Time
(CT): Coagulation Time
(INTEM): Intrinsically activated assays using phospholipid and ellagic acid
(COVID-19): Corona virus disease 2019
(SARS-CoV-2): Severe acute respiratory syndrome coronavirus 2
(ICU): Intensive Care Unit
(CCT): Conventional coagulation test
(PCR): Polymerase chain reaction
(CT): Computed tomography
(NIV): Non-invasive ventilation
(NHF): Nasal high flow oxygen therapy
(COPD): Chronic obstructive pulmonary disease
(INR): International Normalized Ratio
(LMWH): Low Molecular Weight Heparin
(FIBTEM): Fibrin-based extrinsically activated assays with tissue factor and platelet inhibitor cytochalasin D
(HEPTEM): Intrinsically activated assays with the addition of heparinase
(CFT): Clot Formation Time
(LI): Lysis Index
(BMI): Body mass index
(AUC): Area under the curve
(ROC): Receiver operating characteristic
(IQR): Interquartile range
(STROBE): Strengthening the Reporting of Observational Studies in Epidemiology
(DOAC): Direct oral anticoagulants
(RF): Respiratory frequency
(PAI-1): Plasminogen activator inhibitor type 1

## Declarations

### Ethics approval and consent to participate

The study was approved by the Swedish Ethical Review Authority (D-nr 2020-01875).

## Acknowledgements

We wish to thank the staff at the COVID-19 wards at Capio St Göran’s Hospital for participation and collaboration, the staff at the Laboratory Unit (especially Jacqueline Akcan) for conducting the blood samples and colleagues at the Intensive Care Unit for support. Special thanks to Rasmus Berglind, Anton Borgström and Christine Carlswärd for valuable assistance with data collection.

